# Implementation of a Multimodal Diagnostic Algorithm for Blood Culture-Negative Infective Endocarditis at the Argentine National Reference Laboratory: A Prospective Study

**DOI:** 10.64898/2026.08.06.26359889

**Authors:** R Armitano, G Martinez, M Prieto

## Abstract

**Background:** Blood culture-negative infective endocarditis (BCNIE) poses a significant diagnostic challenge. This study evaluated a multimodal diagnostic algorithm combining serological and molecular methods at the Argentine National Reference Laboratory.

**Methods:** A prospective analysis was conducted on 53 consecutive patients with suspected BCNIE referred between January 2019 and December 2024. The diagnostic workflow included indirect immunofluorescence for *Bartonella* spp. and *Coxiella burnetii*, species-specific PCR for *Bartonella* spp. and *Tropheryma whipplei*, and broad-range 16S rRNA PCR with Sanger sequencing on available blood and valvular tissue specimens.

**Results:** An etiological diagnosis was established in 17 of 53 patients (32.1%). *Bartonella* spp. was the predominant pathogen (47.1%; 8/17), followed by *T. whipplei* (35.3%; 6/17) and *Streptococcus* spp. (17.6%; 3/17). All *Bartonella* cases were initially detected via serology, with molecular confirmation achieved exclusively through valvular tissue analysis.

**Conclusions:** Implementing a standardized multimodal diagnostic algorithm significantly enhances etiological yields in BCNIE. The findings emphasize the complementary value of frontline serology and targeted molecular testing, highlighting that simultaneous submission of serum, blood, and valvular tissue is essential for optimal diagnosis.

## Introduction

Infective endocarditis (IE) remains a severe, life-threatening condition associated with high morbidity and mortality, wherein the early identification of the etiological agent is essential to establish targeted antimicrobial therapy and improve patient prognosis (1). While culture-based techniques remain a traditional cornerstone for diagnosis, blood culture-negative infective endocarditis (BCNIE) poses a significant clinical challenge. BCNIE accounts for approximately 5% to 20% (and up to over 30% globally) of all IE cases (2-4). This epidemiological disparity is primarily driven by multiple factors, including differences in diagnostic criteria, regional distribution of fastidious zoonotic pathogens, high rates of prior antimicrobial administration before blood culture collection, variations in sampling and testing strategies, and the presence of non-cultivable bacterial pathogens or non-infectious etiologies (5).

The updated 2023 Duke-International Society for Cardiovascular Infectious Diseases (ISCVID) criteria introduced major modifications to the microbiological diagnosis of IE, placing heightened emphasis on serological testing for *Coxiella burnetii* and *Bartonella* (2).

This study aimed to implement and evaluate, as a pilot experience within a National Reference Laboratory framework, a multimodal diagnostic algorithm combining serological assays and molecular methods to enhance the etiological identification of patients with suspected BCNIE.

## Materials and Methods

### Study population

A prospective study was conducted including consecutive patients with a clinical diagnosis of BCNIE who were referred to the National Reference Laboratory (NRL) between January 2019 and December 2024. The diagnosis of IE and the classification as BCNIE were established by the treating physicians at the referring healthcare institutions according to their routine clinical evaluation. All patients included in the study had persistently negative blood cultures at their respective healthcare centers before referral. A total of 53 patients from healthcare institutions throughout Argentina were included. This study was designed as a pilot evaluation of a laboratory diagnostic algorithm for BCNIE. Consequently, only limited demographic information (age and sex) was available for analysis, and all additional patient information was anonymized before inclusion in the study.

Before sample referral, the NRL provided all participating institutions with standardized written instructions describing the recommended specimens for the diagnostic investigation of suspected BCNIE. These guidelines recommended the simultaneous submission of serum for serological testing, EDTA-anticoagulated whole blood for pathogen-specific molecular assays, and, whenever available, excised heart valve tissue and/or valvular vegetations obtained during cardiac surgery for molecular analysis. However, the specimens ultimately received depended on the clinical management of each patient, specimen availability, and adherence to the referral recommendations at the participating healthcare institutions.

### Diagnostic algorithm

A stepwise diagnostic algorithm was implemented for the etiological investigation of BCNIE. The initial diagnostic approach consisted of serological testing for *Bartonella* spp. and *Coxiella burnetii*. IgG antibody titer ≥1:800 for *Bartonella* spp. or phase I IgG antibody titer ≥1:800 for *C. burnetii* was considered diagnostic, based on extensive evidence from the international literatura (3,5). These serological criteria were subsequently incorporated as major microbiological criteria in the 2023 Duke– International Society for Cardiovascular Infectious Diseases (Duke-ISCVID) diagnostic criteria for infective endocarditis (2). Patients with positive serological results subsequently underwent specific PCR assays for *Bartonella* spp. followed by sequencing to obtain molecular confirmation and species-level identification whenever posible.

In patients with negative serological results, molecular testing was performed according to sample availability. When excised valvular tissue was available, broad-range bacterial 16S rRNA PCR followed by sequencing was performed in parallel with species-specific PCR assays targeting *Bartonella* spp. and *Tropheryma whipplei*. When valvular tissue was unavailable, species-specific PCR assays were performed on EDTA-anticoagulated whole-blood samples.

### Serological testing

Serum samples were analyzed by indirect immunofluorescence assay (IFA) using commercially available kits (DiaSorin, Saluggia, Italy, and Vircell S.L., Granada, Spain) for the detection of IgG antibodies against *Bartonella* spp. and phase I *C. burnetii* antigens, according to the manufacturers’ instructions.

Samples were initially screened at a dilution of 1:64 (*Bartonella* sp.) and 1/16 (*C. burnetii*). Reactive sera were subsequently titrated by twofold serial dilutions till 1:1024 to determine the endpoint antibody titer. When antibody titers were >1:512 but <1:1024, a follow-up serum sample was requested for repeat testing to assess changes in antibody titers. Slides were examined by fluorescence microscopy by experienced laboratory personnel. Samples with antibody titers exceeding the highest dilution tested were reported as 1:1024.

Each assay included the positive and negative controls provided by the manufacturer. Test results were considered valid only when the controls yielded the expected reactivity.

### Molecular analysis

Genomic DNA was extracted from EDTA-anticoagulated whole-blood samples and excised valvular tissue and/or vegetations using the DNeasy Blood & Tissue Kit (Qiagen, Hilden, Germany), according to the manufacturer’s instructions.

Species-specific PCR assays were performed for the detection of *Bartonella* spp. and *T. whipplei. Bartonella* DNA was detected by amplification of the 16S–23S rRNA intergenic spacer (ITS) region using the primers and PCR conditions previously described (6). *T. whipplei* DNA was detected by nested PCR targeting a partial fragment of the *hsp65* gene, following the protocol described by Morgenegg *et al*.(7).

Broad-range bacterial PCR targeting the 16S rRNA gene was performed using the primers and cycling conditions described by Millar *et al*.(6). Amplicons were analyzed by electrophoresis in 2% agarose gels, purified using the QIAquick PCR Purification Kit (Qiagen), and sequenced by the Sanger chain-termination method on an ABI Prism 377 Genetic Analyzer (Applied Biosystems, Foster City, CA, USA). The resulting sequences were compared with reference sequences deposited in the GenBank database (National Center for Biotechnology Information, NCBI) using the BLAST algorithm. Bacterial identification was assigned based on the highest sequence identity (≥99%) and query coverage relative to reference sequences.

To ensure the reliability of molecular results, each analytical run included positive and negative extraction controls as well as positive and negative PCR controls. In addition, amplification of the human β-globin gene was used as control to assess DNA integrity and the presence of PCR inhibitors (6). Molecular results were considered valid only when all quality-control parameters were met. All positive broad-range 16S rRNA and *T. whipplei* PCR amplicons were confirmed by Sanger sequencing to verify the etiological agent and exclude nonspecific amplification or cross-reactivity. Molecular findings were interpreted in conjunction with the available serological and microbiological results.

## Results

The study cohort included 53 patients. The mean age was 46.8 years (range, 9–83 years), and 39 (73.6%) were male. An etiological diagnosis was established in 17 patients (32.1%). *Bartonella* spp. was the most frequently identified pathogen, accounting for 8 of 17 diagnoses (47.1%; 15.1% of the total cohort), followed by *T. whipplei* (6/17, 35.3%; 11.3%) and *Streptococcus* spp. (3/17, 17.6%; 5.7%). Two patients had evidence of *Bartonella* spp./*C. burnetii* co-infection. The distribution of clinical specimens and microbiological findings is summarized in Table 1.

**Table 1.** Clinical specimens, laboratory results, and etiological diagnosis.

| Patient | Serology<br>*Bartonella*<br>spp | Serology<br>*C. burnetii*<br>Fase I | 16S rRNA<br>PCR<br>Sequencing | Specific<br>PCR<sup>1</sup><br>Blood | Specific<br>PCR<sup>1</sup><br>valvular<br>tissue | Etiological diagnosis |
| --- | --- | --- | --- | --- | --- | --- |
| 1 | 1/2048 | < 1/16 | NT | ND | ND | *Bartonella* sp. |
| 2 | 1/1024 | < 1/16 | NA | ND | NA | *Bartonella* sp. |
| 3 | 1/1024 | < 1/16 | NT | ND | NA | *Bartonella* sp. |
| 4 | 1/1024 | < 1/16 | NT | ND | NA | *Bartonella* sp. |
| 5 | 1/1024 | < 1/16 | NT | ND | NA | *Bartonella* sp. |
| 6 | 1/1024 | 1/1024 | ND | ND | Pos | *Bartonella henselae*<br>(*C. burnetii* infection?) |
| 7 | 1/1024 | 1/1024 | ND | ND | NA | *Bartonella* sp-*C. burnetii* co-infection? |
| 8 | 1/1024 | < 1/16 | NT | NA | Pos. | *Bartonella henselae* |
| 9 | NT | NT | NT | Pos. | NA | *Tropheryma whipplei* |
| 10 | < 1/64 | < 1/16 | NT | Pos | Pos | *Tropheryma whipplei* |
| 11 | < 1/64 | < 1/16 | NT | NT | Pos | *Tropheryma whipplei* |
| 12 | < 1/64 | < 1/16 | NT | Pos | NA | *Tropheryma whipplei* |
| 13 | < 1/64 | < 1/16 | NT | Pos | Pos | *Tropheryma whipplei* |
| 14 | NT | NT | NT | NA | Pos | *Tropheryma whipplei* |
| 15 | < 1/64 | < 1/16 | Pos<br>(valvular tissue) | NA | ND | *Streptococcus lutetiensis* |
| 16 | < 1/64 | < 1/16 | Pos<br>(valvular tissue) | Neg | Neg. | *Streptococcus sanguinis* |
| 17 | NT | NT | Pos<br>(valvular tissue) | NA | ND | *Streptococcus* sp. |

<sup>1</sup>*Bartonella* sp, *T. whipplei*
NT, Not tested ; NA, Not clinical specimen available , ND: No DNA was detected; Pos., DNA was a

## Discussion

This study demonstrates that implementation of a standardized multimodal diagnostic algorithm substantially improves the microbiological diagnosis of BCNIE in a NRL setting. An etiological diagnosis was established in nearly one-third of patients, underscoring the complementary value of serology, pathogen-specific PCR, and broad-range 16S rRNA sequencing.

An important finding of this study is that 15 of the 17 patients with a confirmed etiological diagnosis had serum samples submitted in addition to EDTA blood and/or valvular tissue. Notably, all cases of *Bartonella* IE were initially identified by serology, with results available within 24 hours of sample receipt. In contrast, *Bartonella* DNA could not be detected by species-specific PCR in peripheral blood. Molecular confirmation was achieved exclusively in patients for whom valvular tissue was available. These findings indicate the poor diagnostic yield of peripheral blood PCR for *Bartonella* endocarditis and reinforce the central role of serology as the first-line diagnostic approach for BCNIE. Given its rapid turnaround time, low cost, and wide availability, serology represents a practical and accessible diagnostic tool that can be readily incorporated into routine clinical microbiology laboratories, particularly in settings where cardiac tissue is unavailable.

A noteworthy finding was the unexpectedly high proportion of *T. whipplei* infections, which accounted for more than one-third of all confirmed etiological diagnoses. Although Whipple endocarditis has traditionally been considered a rare cause of BCNIE (8), increasing use of molecular diagnostic techniques has demonstrated that it is probably underrecognized rather than truly uncommon. Unlike *Bartonella* spp. and *C. burnetii*, no validated serological assay is available for routine diagnosis of *T. whipplei* infective endocarditis, making molecular methods essential. Our findings support the incorporation of *T. whipplei*-specific PCR into the routine diagnostic work-up of BCNIE, particularly in referral laboratories.

The distribution of pathogens identified in our cohort differed substantially from that reported in previous large series of BCNIE. While *C. burnetii* has consistently been documented as the leading global cause of true BCNIE, accounting for approximately 28% to 37% of cases, followed by *Bartonella* spp. (12% to 28%) (8); *Bartonella* spp. (47.1%) and *T. whipplei* (35.3%) were the predominant pathogens in our study.

The diagnostic yield depended heavily on the specimens submitted. Nine patients submitted only serum, preventing molecular investigation for *T. whipplei*, whereas five submitted only EDTA blood and/or valvular tissue, precluding serological investigation for *Bartonella* spp. and *C. burnetii*. These observations emphasize that simultaneous submission of serum together with EDTA blood and, whenever available, excised valvular tissue is essential.

Several methodological and operational limitations must be acknowledged. First, the prospective nature of this pilot evaluation was constrained by incomplete sample collection and referral heterogeneity; notably, valvular tissue was unavailable in a subset of cases where clinicians submitted solely peripheral blood or serum samples, which inherently restricted the sensitivity of molecular assays that perform optimally on localized foci of infection. Second, conventional endpoint PCR assays present inherent limitations regarding turnaround time, risk of carryover contamination during post-amplification gel analysis, and the inability to simultaneously quantify bacterial load or process high-throughput sample volumes efficiently. Nevertheless, an essential strength of the current stepwise framework is its cost-effectiveness and feasibility for decentralized implementation. While advanced national reference centers transition toward high-throughput real-time platforms, the strategic combination of accessible serological screening and standard molecular assays provides a robust, economically sustainable model. This approach can be readily adopted by regional healthcare centers to decentralize diagnostics, avoid sample shipping delays, and reduce total reliance on centralized reference laboratories. Looking forward, future directions will focus on translating these workflows into multiplex real-time PCR (qPCR) assays to maximize analytical sensitivity, provide precise quantification, reduce turnaround times, and integrate internal amplification controls. Moreover, expanding the diagnostic panel beyond *Bartonella* spp. and *T. whipplei* to include other fastidious endocarditis-associated pathogens, such as members of the HACEK group (*Haemophilus, Aggregatibacter, Cardiobacterium, Eikenella, Kingella*), *Coxiella burnetii* and *Mycoplasma* species, will ensure comprehensive coverage of the etiological spectrum observed in Latin America. Institutional reinforcement and stepped-care validation of these diagnostic algorithms will ultimately empower regional networks, accelerate targeted antimicrobial therapy, and improve clinical outcomes for BCNIE patients nationwide.

## Data Availability

All data produced in the present work are contained in the manuscript

## Ethics statement

Clinical specimens included in this study were submitted to the National Reference Laboratory (NRL) for diagnostic testing as part of its national reference activities for the investigation of suspected blood culture-negative infective endocarditis. Clinical data were analyzed in an anonymized manner, ensuring patient confidentiality in accordance with applicable national regulations and the ethical principles of the Declaration of Helsinki. As the study used anonymized clinical specimens collected during routine diagnostic practice, no additional informed consent was required.

## References

1. Rajani, R., & Klein, J. L. (2020). Infective endocarditis: A contemporary update. Clinical Medicine, 20(1), 31–35. 10.7861/clinmed.cme.20.1.1

2. Fowler, V. G., Durack, D. T., Selton-Suty, C., Athan, E., Bayer, A. S., Chamis, A. L., Dahl, A., DiBernardo, L., Durante-Mangoni, E., Duval, X., Fortes, C. Q., Fosbøl, E., Hannan, M. M., Hasse, B., Hoen, B., Karchmer, A. W., Mestres, C. A., Petti, C. A., Pizzi, M. N., Preston, S. D., … Miro, J. M. (2023). The 2023 Duke-International Society for Cardiovascular Infectious Diseases Criteria for Infective Endocarditis: Updating the Modified Duke Criteria. Clinical Infectious Diseases, 77(4), 518–526. 10.1093/cid/ciad271

3. Lin, K. P., Yeh, T. K., Chuang, Y. C., Wang, L. A., Fu, Y. C., & Liu, P. Y. (2023). Blood Culture Negative Endocarditis: A Review of Laboratory Diagnostic Approaches. International Journal of General Medicine, 16, 317–327. 10.2147/IJGM.S393329

4. DeSimone, D. C., Garrigos, Z. E., Marx, G. E., Tattevin, P., Hasse, B., McCormick, D. W., Hannan, M. M., Zuhlke, L. J., Radke, C. S., Baddour, L. M., & American Heart Association Council on Lifelong Congenital Heart Disease and Heart Health in the Young; Council on Clinical Cardiology; and Council on Quality of Care and Outcomes Research. (2025). Blood Culture-Negative Endocarditis: A Scientific Statement From the American Heart Association: Endorsed by the International Society for Cardiovascular Infectious Diseases. Journal of the American Heart Association, 14(8), e040218. 10.1161/JAHA.124.040218

5. Fournier, P. E., Gouriet, F., Casalta, J. P., Lepidi, H., Chaudet, H., Thuny, F., Collart, F., Habib, G., & Raoult, D. (2017). Blood culture-negative endocarditis: Improving the diagnostic yield using new diagnostic tools. Medicine, 96(47), e8392. 10.1097/MD.0000000000008392

6. Millar, B., Moore, J., Mallon, P., Xu, J., Crowe, M., Mcclurg, R., Raoult, D., Earle, J., Hone, R., & Murphy, P. (2001). Molecular diagnosis of infective endocarditis--a new Duke’s criterion. Scandinavian Journal of Infectious Diseases, 33(9), 673–680. 10.1080/00365540110026764

7. Morgenegg, S., Dutly, F., & Altwegg, M. (2000). Cloning and sequencing of a part of the heat shock protein 65 gene (hsp65) of “Tropheryma whippelii” and its use for detection of “T. whippelii” in clinical specimens by PCR. Journal of Clinical Microbiology, 38(6), 2248–2253. 10.1128/JCM.38.6.2248-2253.2000

8. Godfrey, R., Curtis, S., Schilling, W. H., & James, P. R. (2020). Blood culture negative endocarditis in the modern era of 16S rRNA sequencing. Clinical Medicine, 20(4), 412–416. 10.7861/clinmed.2019-0342

